# Xylazine Prevalence and Concentration in the Los Angeles Fentanyl Market, 2023 Q1 – 2025 Q2

**DOI:** 10.1101/2025.05.13.25327478

**Authors:** Joseph Friedman, Caitlin A. Molina, Adam J. Koncsol, Ruby Romero, Morgan E. Godvin, Elham Jalayer, spider davila, Oscar Arellano, Amanda Cowan, Brian Hurley, Chelsea L. Shover

**Author notes:** Correspondence to Joseph R. Friedman, MD, PhD, MPH, Department of Psychiatry, University of California, San Diego, 200 W Arbor Dr., San Diego, CA, 92103.

## Abstract

**Background:** The veterinary sedative xylazine has been mostly described on the East Coast—yet early reports indicate that it is now arriving to West Coast fentanyl markets. Emerging drug checking approaches can provide information about the concentration and prevalence of xylazine in illicit fentanyl.

**Methods:** Fentanyl samples from a community-based drug checking program in Los Angeles, California were assessed using direct analysis in real time mass spectrometry (DART-MS). A subset was analyzed with liquid chromatography gas spectrometry (LC-MS) to quantify the concentration of xylazine, fentanyl, and other compounds.

**Results:** Among n=536 fentanyl-positive samples, xylazine positivity rose from 0% in 2023 Q1 to a peak of 29.5% in 2025 Q1. A significant time trend was observed (OR per quarter year= 1.35 [95%CI: 1.19-1.52]). Xylazine concentration in fentanyl samples was generally low, with a highly skewed distribution (mean=2.42%, sd=7.80%). 76.9% of xylazine-positive samples had <1.0% xylazine concentration. Compared to xylazine-negative samples, xylazine-positive samples were more likely to contain BTMPS (46.60% vs 17.30%), and lidocaine (65.0% vs 29.6%), and had lower average fentanyl concentration (6.12% vs 10.7%).

**Conclusions:** Among illicit fentanyl samples in LA, we note increasing xylazine positivity. The distribution of xylazine concentration is highly skewed, with a small number of very high concentration samples, and majority with <1%. Nevertheless, more research is needed to study the health impacts of even the low concentration xylazine that is most predominant here; given that the average participant in our sample consumes about 1.0g of illicit fentanyl daily, the corresponding dose of 24mg of xylazine per day may be physiologically significant.

## Background

The rise of the veterinary sedative xylazine as a fentanyl additive has been linked to more complicated overdose response, worsening soft tissue infections, and other health risks for people who use drugs (Bufanda et al., 2025; Cano et al., 2024; Friedman et al., 2022; Jawa et al., 2024). Consequently, in 2023 xylazine was identified as an emerging threat by the Biden Administration in the United States, and it has been the focus of a growing body of literature (Gupta Rahul et al., 2023; Zagorski et al., 2023).

Xylazine as an illicit drug supply adulterant has been mostly studied—and has been most prevalent—on the East Coast, yet early reports indicate that it is now spreading across the country, and arriving to West Coast fentanyl markets (Bufanda et al., 2025; Friedman, 2024). Xylazine has mostly been characterized in terms of binary presence/absence, either among overdose autopsy toxicology (Cano et al., 2024; Friedman, 2024) or via xylazine testing strips on urine or drug samples (Bowles et al., 2021; Copeland et al., 2024; Hauschild et al., 2023). However, there is great potential value in quantifying the percent concentration of xylazine in illicit fentanyl samples, for improved estimation of exposure among people who use drugs. This is now possible with emerging drug checking approaches. Here we describe both the qualitative prevalence, and quantitative percent concentration, of xylazine among illicit fentanyl samples from a community-based drug checking program in Los Angeles, CA from the first quarter (Q1) 2023 to the second quarter (Q2) of 2025.

## Methods

Samples of drug product were provided anonymously by clients seeking services at a community-based drug checking program in Los Angeles, California. Samples were sent to the National Institute of Standards and Technology (NIST) for laboratory-based qualitative testing using direct analysis in real time mass spectrometry (DART-MS). A subset also underwent quantitative analysis with liquid-chromatography mass spectrometry (LC-MS).

The laboratory methodologies employed here have been previously described (Appley et al., 2022; Sisco et al., 2017). Briefly, DART-MS spectra were analyzed against libraries of over 1,300 substances, including pharmaceutical and illicit drugs, adulterants, cutting and bulking agents, precursor chemicals, and other substances (e.g., adhesives, food products, etc.). The LC-MS quantification panel included xylazine, fentanyl, fentanyl precursor chemicals, other illicit drugs, and common adulterants.

Among fentanyl-positive samples, bivariate logistic regression analysis was used to assess the statistical significance of the xylazine-positivity time trend observed during the study period. Separate bivariate logistic regressions were also employed to assess the relationship between xylazine positivity (predictor) and the presence/absence of BTMPS, heroin, methamphetamine, cocaine, and lidocaine (one regression per outcome variable). Differences in average fentanyl concentration between xylazine positive and negative samples were assessed using a bivariate quasi-Poisson regression, with fentanyl concentration as the outcome variable, and xylazine presence as the predictor. The UCLA Institutional Review Board reviewed and approved this project (protocol IRB-22-0760) and additionally determined that aspects of this work constituted public health surveillance and not human subjects research.

## Results

Among a total of n=536 fentanyl-positive samples analyzed between 2023 Q1-2025 Q2, xylazine positivity rose from 0% of n=17 samples in 2023 Q1 to a peak of 29.5% of n=78 samples in 2025 Q1, before declining slightly in 2025 Q2 to 22.2% of n=45 samples (Figure 1). In bivariate logistic regression analysis, a positive time trend was observed, with each additional quarter year associated with increased odds of xylazine positivity (OR= 1.35 [95%CI: 1.19-1.52]).

**Figure 1.**
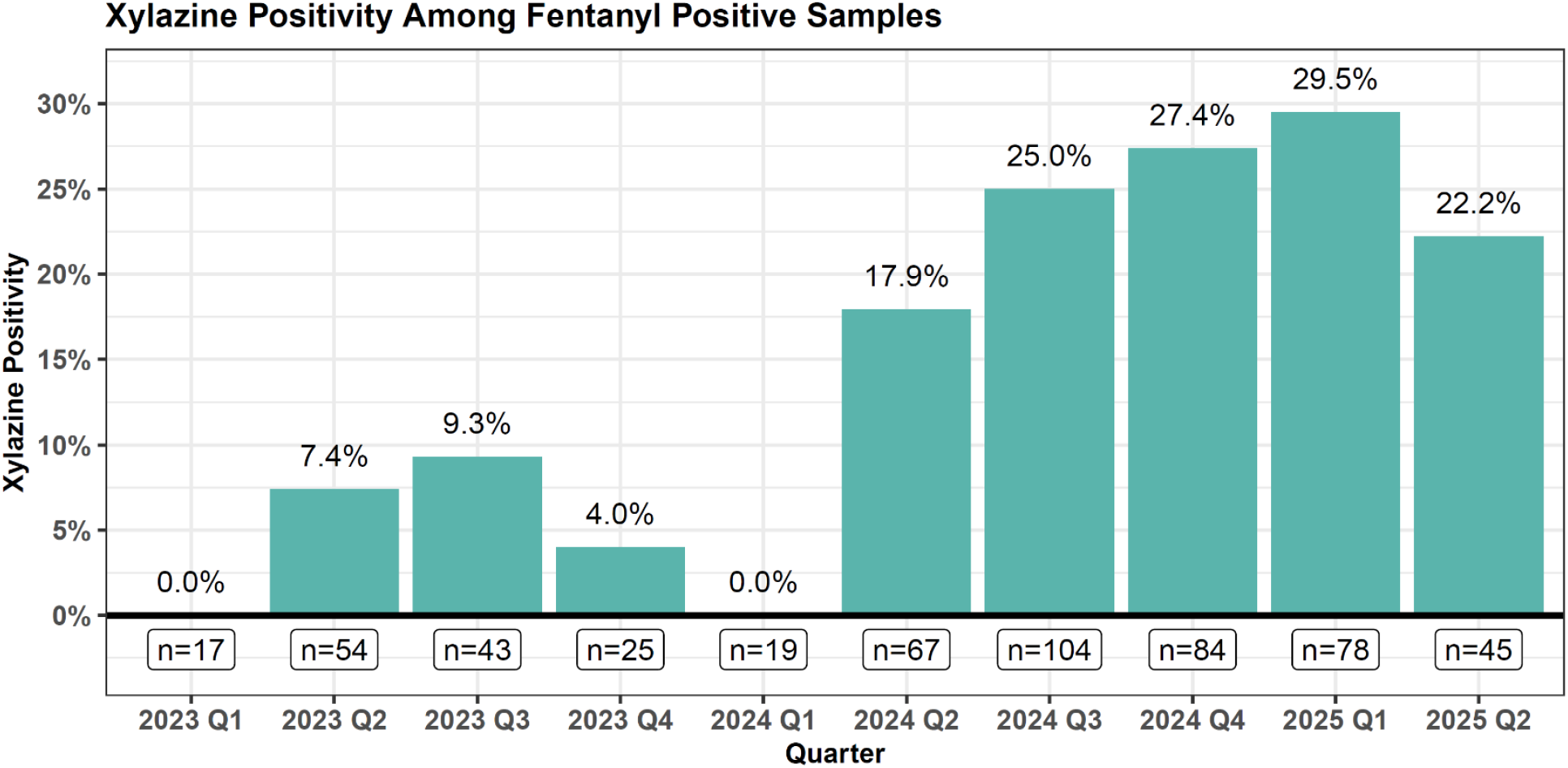
Xylazine Positivity Among Fentanyl-Positive Samples, 2023-2025Q1. Xylazine positivity among fentanyl-positive samples brought to drug checking services in Los Angeles are shown by quarter, between the first quarter (Q1) of 2023 and the second quarter (Q2) of 2025.

Among n=103 xylazine-positive samples, n=78 had quantitative results available. Of these xylazine concentration was generally low, with a highly skewed distribution (mean=2.42, sd=7.80) [Table 1]. 76.9% of xylazine-positive samples had less than 1% xylazine concentration.

**Table 1.** Characteristics of Xylazine Positive and Negative Fentanyl Samples. Characteristics of xylazine-positive and negative samples are compared, including the mean xylazine and fentanyl concentrations, and the co-prevalence of other substances. Differences in fentanyl concentration are assessed using a rate ratio (RR) from a bivariate quasi-Poisson generalized linear model, while differences in substance co-prevalence were assessed using odds ratios (OR) from bivariate logistic regressions.

|  | Xylazine Absent<br>(N=433) | Xylazine Present<br>(N=103) | Overall<br>(N=536) | OR/RR (95%CI) |
| --- | --- | --- | --- | --- |
| **Xylazine Concentration** |  |  |  |  |
| Mean (SD) | --- | 2.42 (7.80) | 2.42 (7.80) | -- |
| Fraction under 1% concentration |  | 0.769 | 0.769 | -- |
| **Fentanyl Concentration** |  |  |  |  |
| Mean (SD) | 10.7 (12.3) | 6.12 (7.23) | 9.55 (11.4) | 0.57 (0.40-0.80) |
| **Other Substance Prevalence (%)** |  |  |  |  |
| BTMPS | 17.30% | 46.60% | 22.90% | 4.17 (2.63-6.60) |
| Cocaine | 5.54% | 3.88% | 5.22% | 0.68 (0.23-2.02) |
| Methamphetamine | 9.70% | 11.70% | 10.10% | 1.23 (0.62-2.43) |
| Heroin | 11.10% | 5.83% | 10.10% | 0.49 (0.20-1.19) |
| Lidocaine | 29.60% | 65.00% | 36.40% | 4.43 (2.82-6.99) |

Xylazine-positive samples had a significantly lower concentration of fentanyl compared to xylazine-negative samples, with a mean concentration of 6.12% (sd=7.23%) vs 10.7% (sd=12.3%) respectively, and an estimated quasi-Poisson rate ratio of 0.57 (95%CI: 0.40-0.80). This indicates that among fentanyl samples, the fentanyl concentration of xylazine-positive samples was on average 43% lower relative to those without the co-detection of xylazine.

Compared to xylazine-negative samples, xylazine-positive samples were more likely to contain BTMPS (46.60% vs 17.30%, OR=4.17; 95%CI: 2.63-6.60), and lidocaine (65.0% vs 29.6%, OR=4.43; 95%CI: 2.82-6.99). No significant differences were observed for heroin, cocaine, or methamphetamine positivity (Table 1).

## Discussion

Among illicit fentanyl samples from community drug checking sites in LA, we note a significant increase in xylazine positivity over time, especially after 2024 Q2. In the most recent data in our study, xylazine was present in 1 out of every 4 to 5 fentanyl samples. This adds to a growing body of literature showing that xylazine is spreading across the country from East to West, and that it has now arrived to the West Coast (Bufanda et al., 2025; Friedman, 2024; Gupta Rahul et al., 2023).

To our knowledge, this also represents the largest study, to-date, of quantitative xylazine testing results. One prior analysis quantified xylazine among n=8 fentanyl samples in Rhode Island, finding that concentration ranged from 0.4%-11.8%. Here, we observed a highly skewed distribution of xylazine concentration, with a small number of high concentration samples, and a majority with concentrations under 1%.

Further study will be required to assess the physiological importance of the lower concentration xylazine most common here, which was generally found at lower concentrations than fentanyl among the same samples. It is not clear if the potential negative health effects associated with xylazine— such as persistent soft tissue infections and injury, cardiovascular complications, and overdose management complicated by synergism with opioids—would be found with lower dose xylazine, or only with the higher doses that comprised the minority of our sample.

Given that the average participant at our study sites consumes about 1.0g of illicit fentanyl daily (Shover, 2025), and the 2.4% average concentration that we observed here, the resulting average dose of 24mg of xylazine daily (among the fraction of participants consuming only xylazine-positive samples) may still be pharmacologically significant. Xylazine has not been extensively studied in humans. Dosing for veterinary anesthetic purposes varies by species, but generally ranges from 0.01–2.5 mg/kg(“Anaesthesia of the dog,” 2014; Valverde and Doherty, 2008). Hypotension and bradycardia have been described in human overdoses with doses as low as 0.73mg/kg (Ruiz-Colón et al., 2014). Assuming an average human adult size of 70kg, the average dose we observe would correspond to 0.34 mg/kg—which does fall in the low end of the range of anesthetic dosing observed in veterinary literature. Furthermore, it is unknown to what degree smoking – a dominant route of administration for illicitly manufactured fentanyl in Los Angeles and the West Coast generally – rather than injecting impacts the bioavailability and health effects of xylazine (Eger et al., 2024).

Additional research will also be required to assess if this distribution of xylazine concentration holds true in other fentanyl markets, or if it is unique to Los Angeles. Given the nature of community-based drug checking, the sample presented here may suffer from convenience bias and is therefore limited in its ability to generalize to the greater West Coast. Other limitations may include that the LC-MS quantitation panel used during the majority of the study window included only twelve substances, and that DART-MS analysis may be unable to detect certain larger-molecule substances such as sugars and other common fillers.

The lower concentration of fentanyl among xylazine-positive samples is also notable. This may indicate that xylazine represents an alternative strategy for achieving a physiologically potent product, compared to higher-concentration fentanyl formulations. Xylazine also had a greater chance of co-occurring with BTMPS, and lidocaine, which are both novel fentanyl adulterants of unclear physiological significance in the context of illicit drug use (Friedman et al., 2024; Shover et al., 2024).

In sum, this work presents one of the first analyses documenting the proliferation of xylazine in the West Coast illicit fentanyl supply of the United States, finding a significantly increasing prevalence over time between 2023 and 2025. It also provides the largest sample of quantitative xylazine testing results to-date. Future research is needed to assess the potential health impacts of low concentration xylazine adulteration of fentanyl samples.

## Data Availability

The underlying data used in this article are highly sensitive and cannot be shared, but authors may be contacted to request summary statistics.

## Acknowledgements

The authors report no conflicts of interest. JRF received funding from the National Institute on Drug Abuse (DA049644) and the National institute of Mental Health (MH101072). CLS received support from the National Institute on Drug Abuse (K01DA050771). AJK received educational support from the NIH/National Center for Advancing Translational Science (NCATS) UCLA CTSI (TL1TR001883). This work was supported by the Centers for Disease Control and Prevention as part of Overdose Data to Action: LOCAL (CDC-RFA-CE-23-0003), and made possible through an equipment grant from the James B. Pendleton Charitable Trust to the UCLA AIDS Institute and UCLA Center for AIDS Research. The funders played no role in the design and conduct of the study; collection, management, analysis, and interpretation of the data; preparation, review, or approval of the manuscript; and decision to submit the manuscript for publication.

## Notes

### Competing Interest Statement

The authors have declared no competing interest.

### Author Declarations

The UCLA Institutional Review Board reviewed and approved this project (protocol IRB-22-0760) and additionally determined that aspects of this work constituted public health surveillance and not human subjects research.

